# Estimation of State Variables and Model Parameters for the Evolution of COVID-19 in the City of Rio de Janeiro

**DOI:** 10.1101/2020.05.21.20108407

**Authors:** Helcio R. B. Orlande, Marcelo J. Colaço, George S. Dulikravich, Luiz Ferreira

## Abstract

**SUMMARY:**

- Evolution model is based on that used by Hernandez et al. [1], which considers the following groups: Susceptible, Incubating, Asymptomatic, Symptomatic, Hospitalized, Recovered and Accumulated deaths.
- Evolution model considers the possibility of infections from asymptomatic, symptomatic and hospitalized individuals.
- Evolution model considers the possibility that individuals who have recovered from the disease become symptomatic again.
- Observation model accounts for underreport of cases and deaths.
- Observation model accounts for delays in reporting cases and deaths.
- Model parameters were initially estimated with the Markov Chain Monte Carlo (MCMC) method [2,3], by using the data of the city of Rio de Janeiro [4] from February 28, 2020 to April 29, 2020. These estimations were used as initial input values for the solution of the state estimation problem for the city of Rio de Janeiro.
- Algorithm of Liu & West for the Particle Filter [5] was used for the solution of the state estimation problem because it allows the simultaneous estimation of state variables and model parameters.
- State estimation problem was solved with the data of the city of Rio de Janeiro [4], from February 28, 2020 to May 05, 2020.
- Monte Carlo simulations were run for 20 future days, considering uncertainties in the model parameters and state variables. Initial conditions were given by the state variables and corresponding distributions estimated with the particle filter on May 05, 2020. Distributions of the model parameters were also given by the estimations obtained for this date.
- Data of the city of Rio de Janeiro [4], from May 06, 2020 to May 15, 2020, were used for the validation of the solution of the state estimation problem.
- **<u>The present model, with the parameters obtained with the Particle Filter, accurately fits the number of reported cases and the number of reported deaths, for 10 days ahead of the period used for the solution of the state estimation problem.</u>**
- The Ratio of Infected Individuals per Reported Cases was around 15 on May 05, 2020.
- The Indexes of Under-Reported Cases and Deaths were around 12 and 2, respectively, on May 05, 2020.
- The Effective Reproduction Number was around 1.6 on February 28, 2020 and dropped to around 0.9 on May 05, 2020. However, uncertainties related to this parameter are large and the effective reproduction number is between 0.3 and 1.5, at the 95% credibility level.
- **The particle filter must be used to <u>periodically update the estimation of state variables and model parameters, so that future predictions can be made.</u>**
- Day 0 is February 28, 2020.

## EVOLUTION MODEL

The evolution model used in this work is based on that of reference [1], which is illustrated by Figure 1.

**Figure 1.**
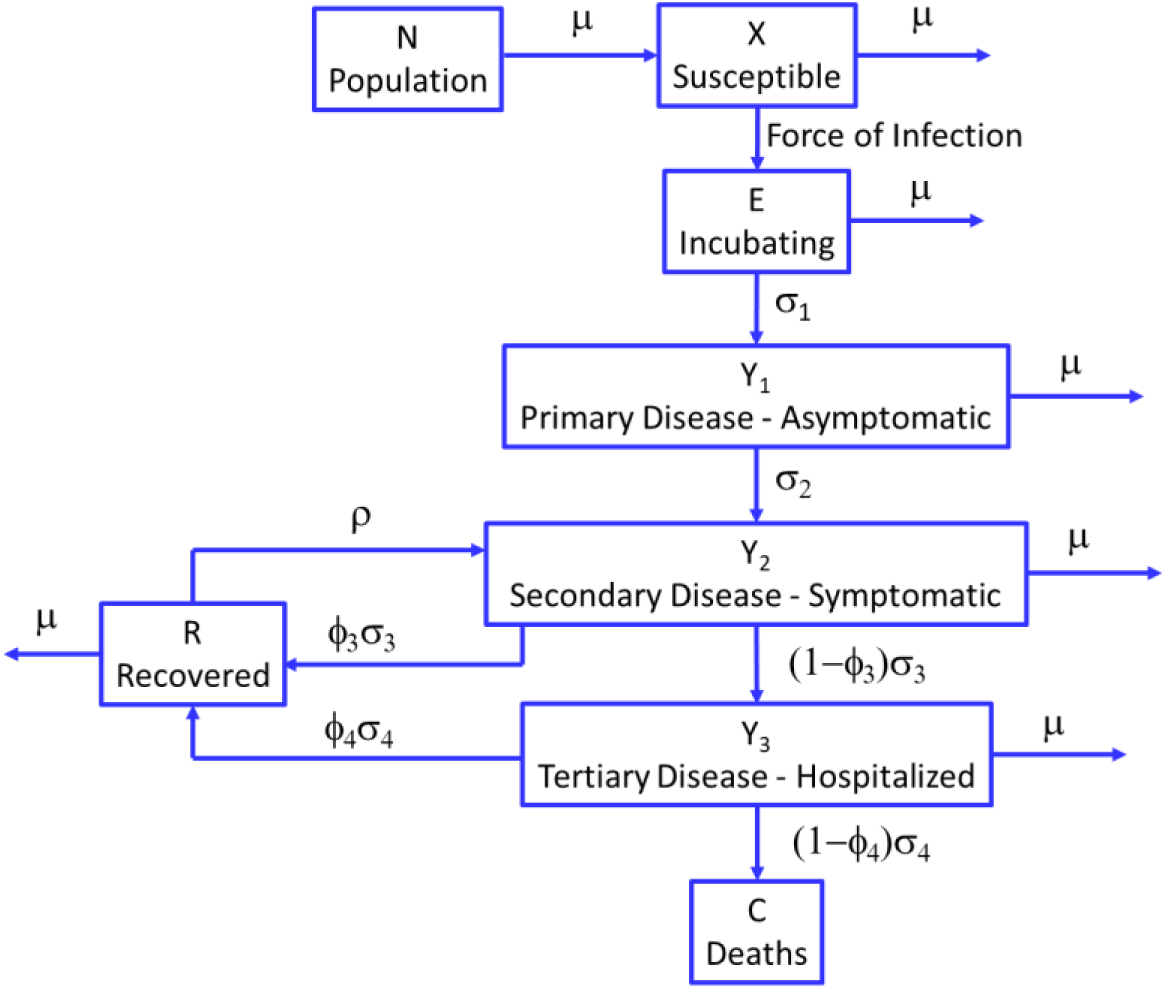
Diagram of the evolution model

The following system of ordinary differential equations represent the model shown in figure 1, for t > 0:

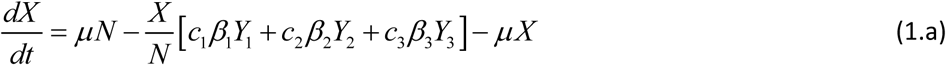

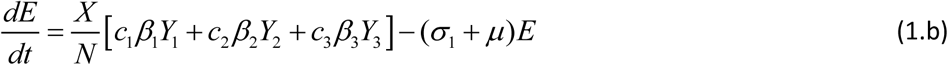

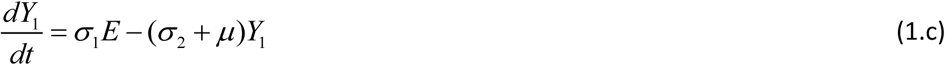

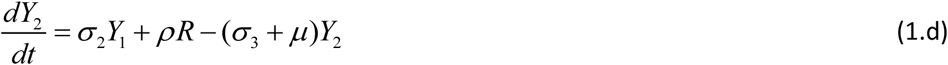

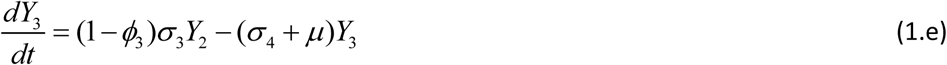

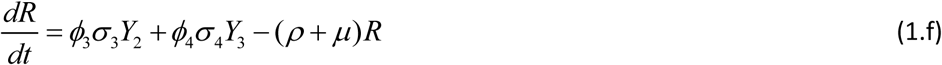

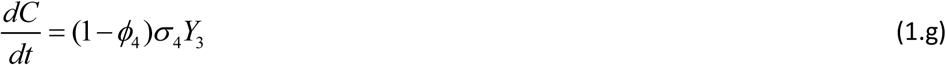

with initial conditions at t = 0 given by:

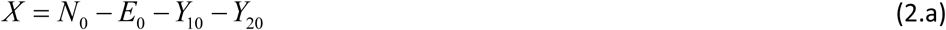

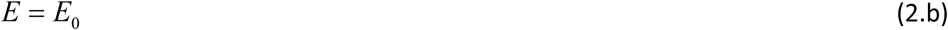

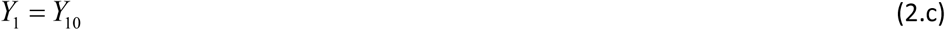

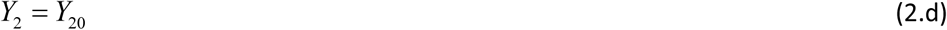

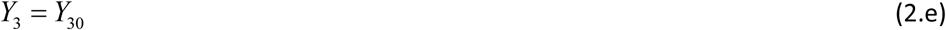

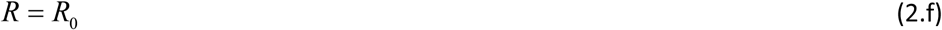

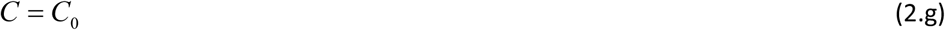

Equations (1.a-g) also need to satisfy the following constraint:

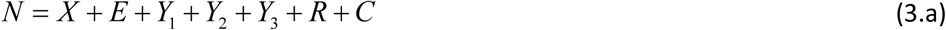

Due to the short time period that the solution of the system (1,2) is sought, constraint (3.a) was substituted by:

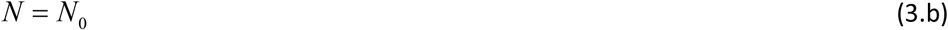

that is, the total population N does not change in time and remains equal to the initial population N_0_.

In the model given above by equations (1-3), the state variables are:

X = number of susceptible individuals

E = number of individuals in the incubation period of the disease

Y_1_ = number of contaminated but asymptomatic individuals

Y_2_ = number of symptomatic individuals

Y_3_ = number of hospitalized individuals

R = number of individuals recovered from the disease

C = accumulated number of deaths

The parameters of the evolution model are as follows:

c_1_ = deterministic parameter to account for infection from asymptomatic individuals

c_2_ = deterministic parameter to account for infection from symptomatic individuals

c_3_ = deterministic parameter to account for infection from hospitalized individuals

β_1_ = transmission probability from asymptomatic individuals

β_2_ = transmission probability from symptomatic individuals

β_3_ = transmission probability from hospitalized individuals

σ_1_ = rate of progression from incubating to asymptomatic individuals

σ_2_ = rate of progression from asymptomatic to symptomatic individuals

σ_3_ = rate of progression from symptomatic to hospitalized individuals

σ_4_ = rate of deaths from hospitalized individuals

ρ = rate of recovered individuals who become symptomatic again

ϕ_3_ = probability of symptomatic individuals to recover

ϕ_4_ = probability of hospitalized individuals to recover

μ = birth rate and death rate by other causes

## MEASUREMENT MODEL

The measurement model considers that the number of reported cases, M_d_, result from the numbers of symptomatic and hospitalized individuals, Y_2_ and Y_3_, respectively. The parameters a_2_ and a_3_ represent the probabilities that individuals, respectively in these two groups, be tested for the disease each day. Delays in reporting the performed tests are accounted for by the rates θ_2_ and θ_3_, for symptomatic and hospitalized individuals, respectively. The diagram presented by figure 2 illustrates the measurement model for the number of reported cases, which is given

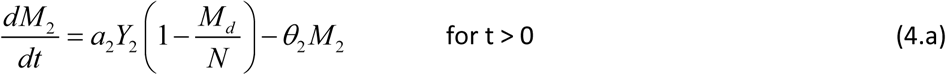

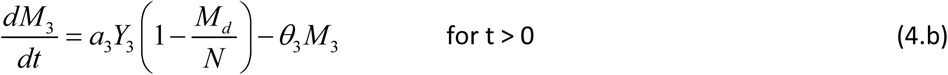

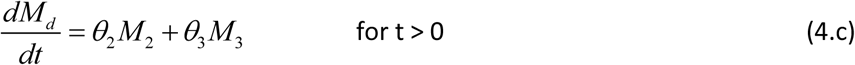

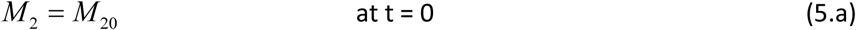

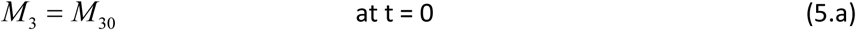

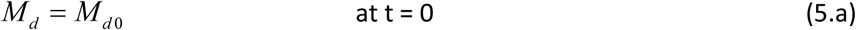

**Figure 2.**
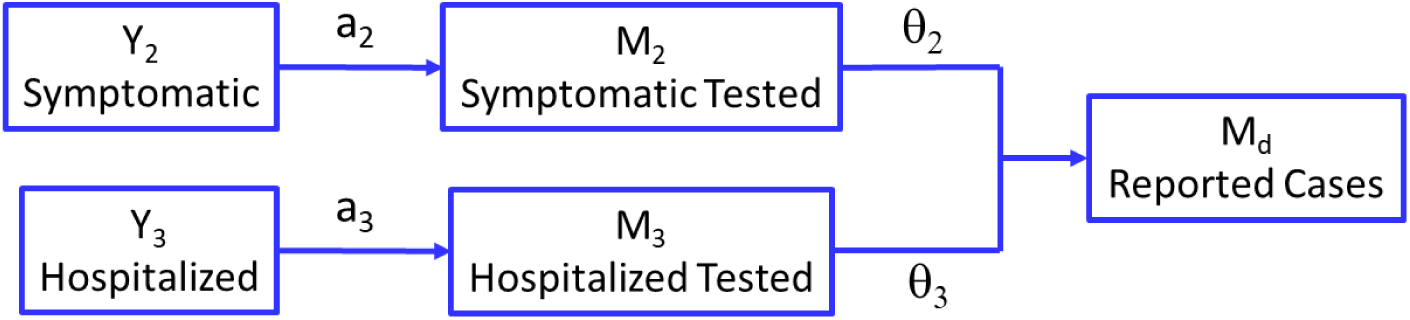
Diagram of the measurement model for the reported cases

The growth rates for symptomatic individuals and hospitalized individuals being tested, M_2_ and M_3_ in equations (4.a) and (4.b), respectively, involves a logistic function, which considers the saturation of the total number of reported cases, M_d_, at the size of the population, N.

The measurement model for reported deaths is analogous to that for reported cases, as shown by figure 3. The equations for the measurement model for reported deaths are given by:

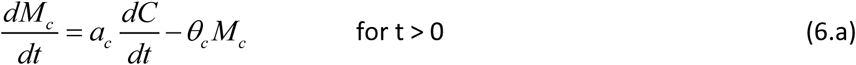

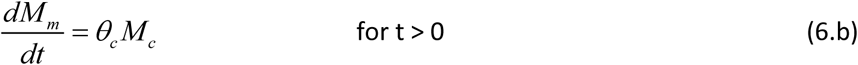

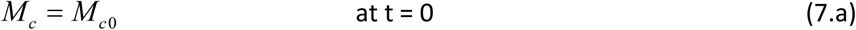

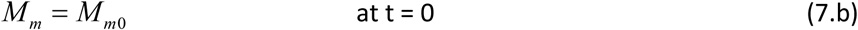

where, in equation (6.a), the rate 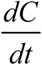 was used instead of the function C because this is an accumulative quantity, which naturally saturates M_c_ when C is saturated. In equations (6.a.b), a_c_ is the probability that any deceased individual be tested for the disease, while θ_c_ is the delay in reporting deaths due to the disease.

**Figure 3.**
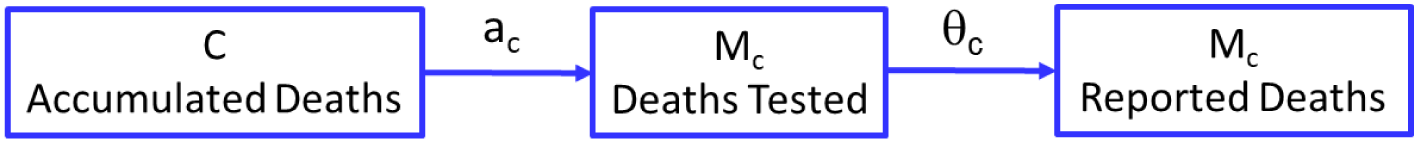
Diagram of the measurement model for the reported deaths

## INITIAL CONDITIONS

The numbers of recovered and hospitalized individuals, as well as deaths and other variables related to the measurement model, were assumed as zero at the date set as the initial time t=0.

In reference [6], an incubation period of 5.1 days was assumed for COVID-19. In order to account for the incubation period, the delay in reporting the first case and existing asymptomatic cases, the simulations were arbitrarily started 7 days before the date of the first reported case. The follow initial conditions were used:

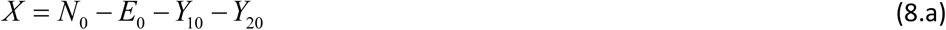

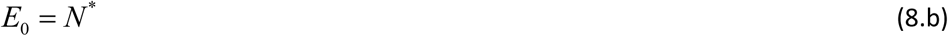

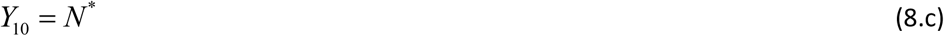

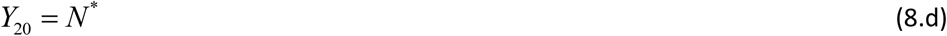

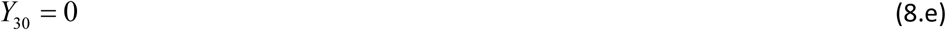

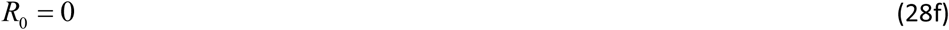

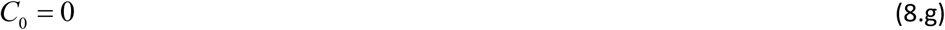

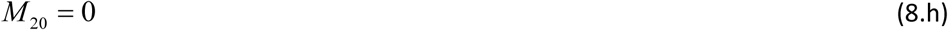

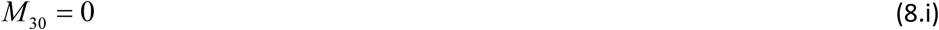

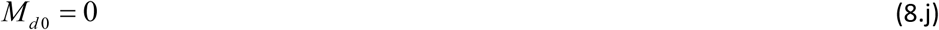

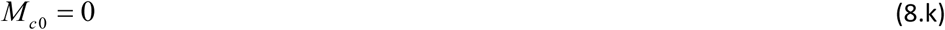

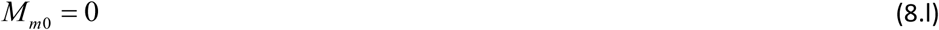

N^*^ was estimated as follows: let 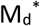 be the first number of cases reported and 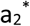 the first guess of the probability of symptomatic individuals be tested. Then, the initially supposed number of infected individuals, 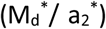, was divided equally in the groups of incubating, asymptomatic and symptomatic individuals, that is,

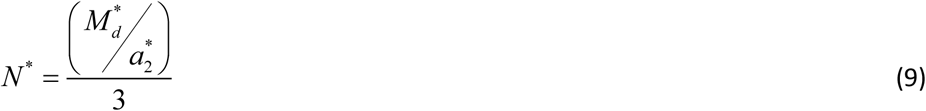

## PARAMETER ESTIMATION USING MCMC

The Metropolis-Hastings algorithm of the Markov chain Monte Carlo (MCMC) method [2,3] was applied for the estimation of the model parameters. This method was used in references [7,8] for parameter estimation in models similar to the present one.

Only the observations (numbers of reported cases and reported deaths) from the city of Rio de Janeiro between February 28, 2020 and May 05, 2020 were used in the parameter estimation with MCMC. Observations were assumed additive, Gaussian, uncorrelated, with zero mean and standard deviations of **10**% of the corresponding observation.

The priors for the parameters P_j_, j = 1,…,N, were assumed as independent Gaussian distributions, truncated to the interval 0 < P_j_ < 1. Efforts were made to use for the means of the Gaussian priors values reported in the literature for COVID-19. The mean values used for the priors are presented in Table 1. In order to account for possible differences of these reported values and the actual ones related to the data of the city of Rio de Janeiro, the standard deviations assigned for the parameters were 15% of the means, except for μ, ϕ_3_ and ϕ_4_. The parameter μ is the birth rate and death rate by causes other than COVID-19. This value has been practically constant in Brazil for more than 10 years. Then, for this parameter, the assigned standard deviation of the prior was 1% of the mean value. The parameters ϕ_3_ and ϕ_4_ represent the probabilities of symptomatic and hospitalized individuals to recover. Mean values reported by different references [6,14] do not differ significantly for these two parameters, which were then assigned with standard deviations of 3% of the mean values for their priors.

The Markov chains were run for 10^5^ states and 7×10^4^ states were considered for the burn-in period. Although convergence was not fully achieved for all chains, the agreement between estimated and observed quantities was very much improved from the initial guess used.

Figures 4a and 4b compare the number of reported cases estimated with the means of the priors and with the last 3×10^4^ states of the Markov chains (yellow lines), respectively, with the observed data. Figures 5a and 5b are analogous to figures 4.a,b, but refer to the number of reported deaths. These figures clearly show the great improvement of the agreement of estimated and observed quantities after the MCMC method was run. We note, however, that such an agreement could not be further improved by considering constant values for the parameters, like in the model presented above. Due to this fact, the particle filter algorithm of Liu & West, which models the parameters as Gaussian mixtures that are dynamically estimated together with the state variables, was used as described in the next section.

**Table 1.** – Means for the Priors

| Parameter | Mean Value | Note |
| --- | --- | --- |
| $c_1$ | 1 | 1 |
| $c_2$ | 1 | 1 |
| $c_3$ | 1 | 1 |
| $\beta_1, \text{day}^{-1}$ | 0.5 | 2 |
| $\beta_2, \text{day}^{-1}$ | 0.5 | 2 |
| $\beta_3, \text{day}^{-1}$ | 0.01 | 3 |
| $\sigma_1, \text{day}^{-1}$ | 1/2.5 | 4 |
| $\sigma_2, \text{day}^{-1}$ | 1/2.5 | 4 |
| $\sigma_3, \text{day}^{-1}$ | 1/10 | 5 |
| $\sigma_4, \text{day}^{-1}$ | 1/12 | 6 |
| $\rho, \text{day}^{-1}$ | $10^{-9}$ | 7 |
| $\phi_3$ | 0.95 | 8 |
| $\phi_4$ | 0.88 | 8 |
| $a_2, \text{day}^{-1}$ | 0.13 | 9 |
| $a_3, \text{day}^{-1}$ | 0.13 | 9 |
| $a_c$ | 0.25 | 10 |
| $\mu, \text{deaths per capita per day}$ | $1.64 \times 10^{-5}$ | 11 |
| $\theta_2$ | 1/5 | 12 |
| $\theta_3$ | 1/5 | 12 |
| $\theta_c$ | 1/4 | 12 |

Note 1: Deterministic value (= 1 means that the group is contagious).
Note 2: Value arbitrarily set in the middle of the feasible interval, due to the lack of more accurate information about the COVID-19 in Brazil. However, we note the following reported values for the SEIR model:
$\beta = \frac{1}{4}$ mean value for Europe [13]
$\beta = 1/1.47$ mean value for China [14]
$\beta = 1/3.38$ mean value for USA [14]
Note 3: Value arbitrarily set, with basis on the qualitative information that the infectious rate of hospitalized individuals is much smaller than those of asymptomatic and symptomatic individuals, because of safety procedures in the hospital.
Note 4: The incubation period of 5.1 days reported in reference [6], was approximately divided for the incubation and primary transmission rates. However, we note the following reported values in reference [14]:
$\sigma_1 = 1/3 \text{ day}^{-1}$ and $\sigma_2 = \frac{1}{2} \text{ day}^{-1}$ .
Note 5: $\sigma_3 = 1/11.3 \text{ day}^{-1}$ estimated in reference [8].
From reference [14]: $\phi_3 = 0.95$ stay symptomatic for 9 days and recover. In 4 days, $(1-\phi_3) = 0.05$ are hospitalized. $9 \phi_3 + 4 (1-\phi_3) = 8.75 \text{ day}$ .
Note 6: $\sigma_4 = 1/10.4 \text{ day}^{-1}$ estimated in reference [6]. $\sigma_4 = 1/14 \text{ day}^{-1}$ reported in reference [14]

**Figure 4.**
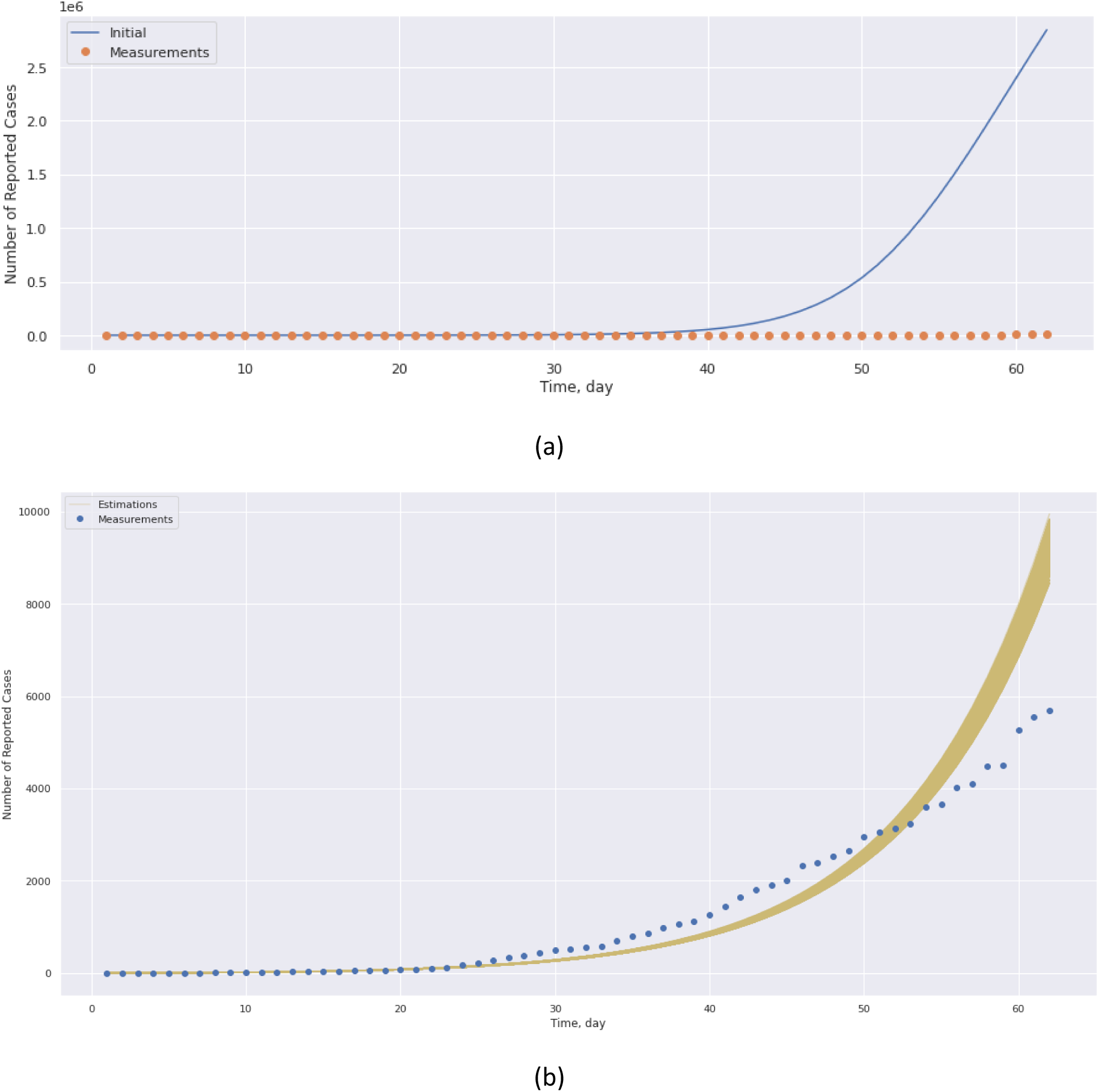
Comparison of estimated and observed number of reported cases obtained with: (a) Mean values of the prior; (b) Simulations with the last 3×10^4^ states of the Markov chains (circles represent the measurements and yellow lines represent the simulations with the parameters of the Markov chains)

**Figure 5.**
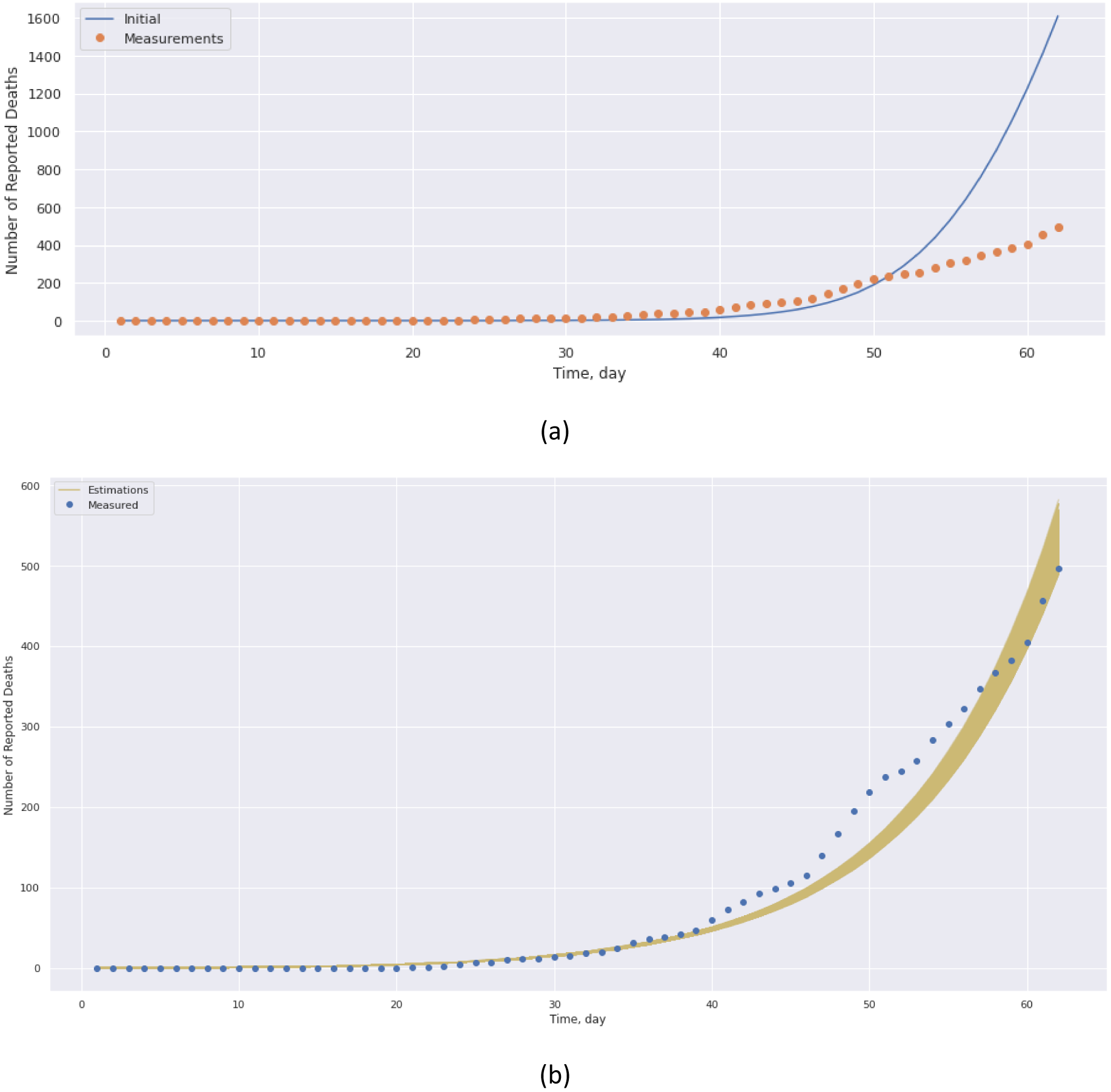
Comparison of estimated and observed number of reported deaths obtained with: (a) Mean values of the prior; (b) Simulations with the last 3×10^4^ states of the Markov chains (circles represent the measurements and yellow lines represent the simulations with the parameters of the Markov chains)

Figures 6 show the histograms of the marginal posterior distributions obtained with the last 3×10^4^ states of the Markov chains for each model parameter. These figures show that some histograms are multimodal or resemble uniform distributions, but several histograms can be fairly approximated by Gaussian distributions. The values of the parameters at the beginning of the Markov chains, as well as the means and the 2.5% and 97.5% quantiles of the last 3×10^4^ states, are presented in these figures. The estimated posterior distributions show that the actual transmission probabilities from asymptomatic and symptomatic individuals, β_1_ and β_2_, are smaller than the values initially supposed. The transmission probability of hospitalized individuals, β_3_, was not significantly modified. As for the progression rates, (σ_1_, σ_2_, σ_3_ and σ_4_), they are all larger than their initial mean values. The estimated probabilities of recovery (ϕ_3_ and ϕ_4_) are significantly smaller than those reported in the literature, which have been used as the initial mean values. The behaviors of the parameters related to the measurement model for the reported cases were as follows: the estimated probability of symptomatic individuals be tested, a_2_, and the rate of reporting the tests, θ_2_, were smaller than the values initially assigned for the means; the analogous parameters for the tests of hospitalized individuals remained unchanged, although θ_3_ exhibits a reduction trend. For the measurement model of deaths, both the test probability (a_c_) and the report rate (θ_c_) increased.

**Figure 6.**
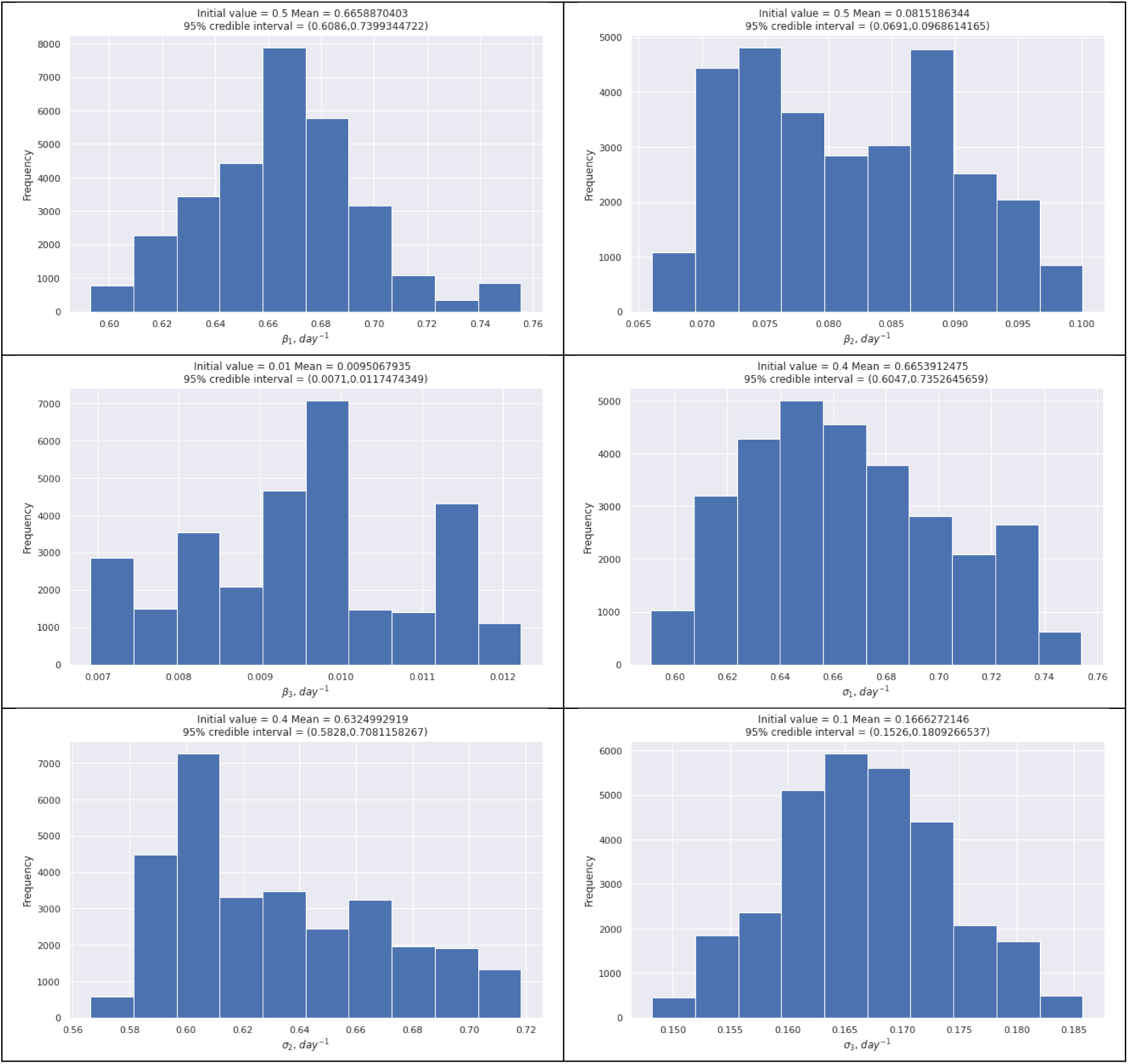
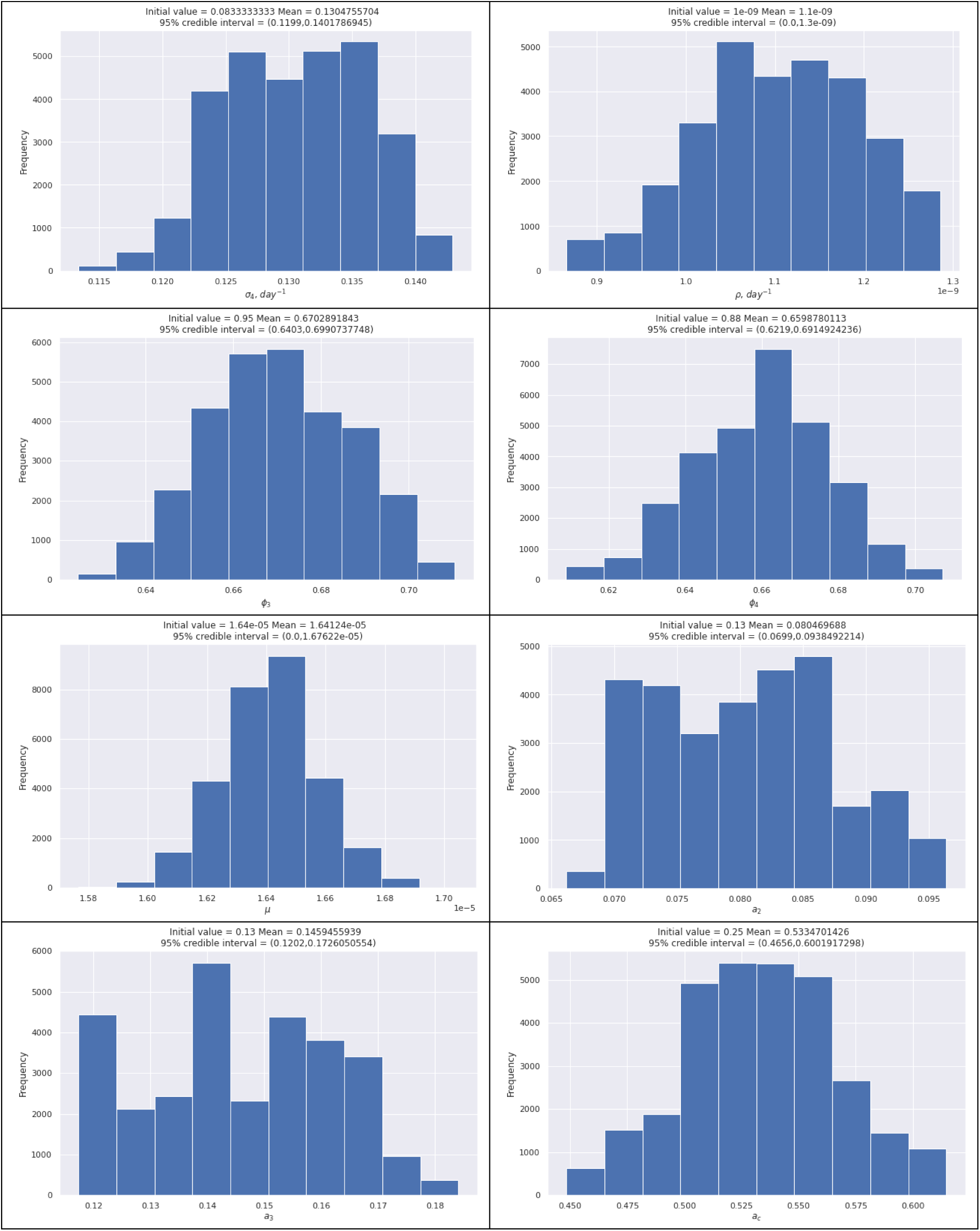
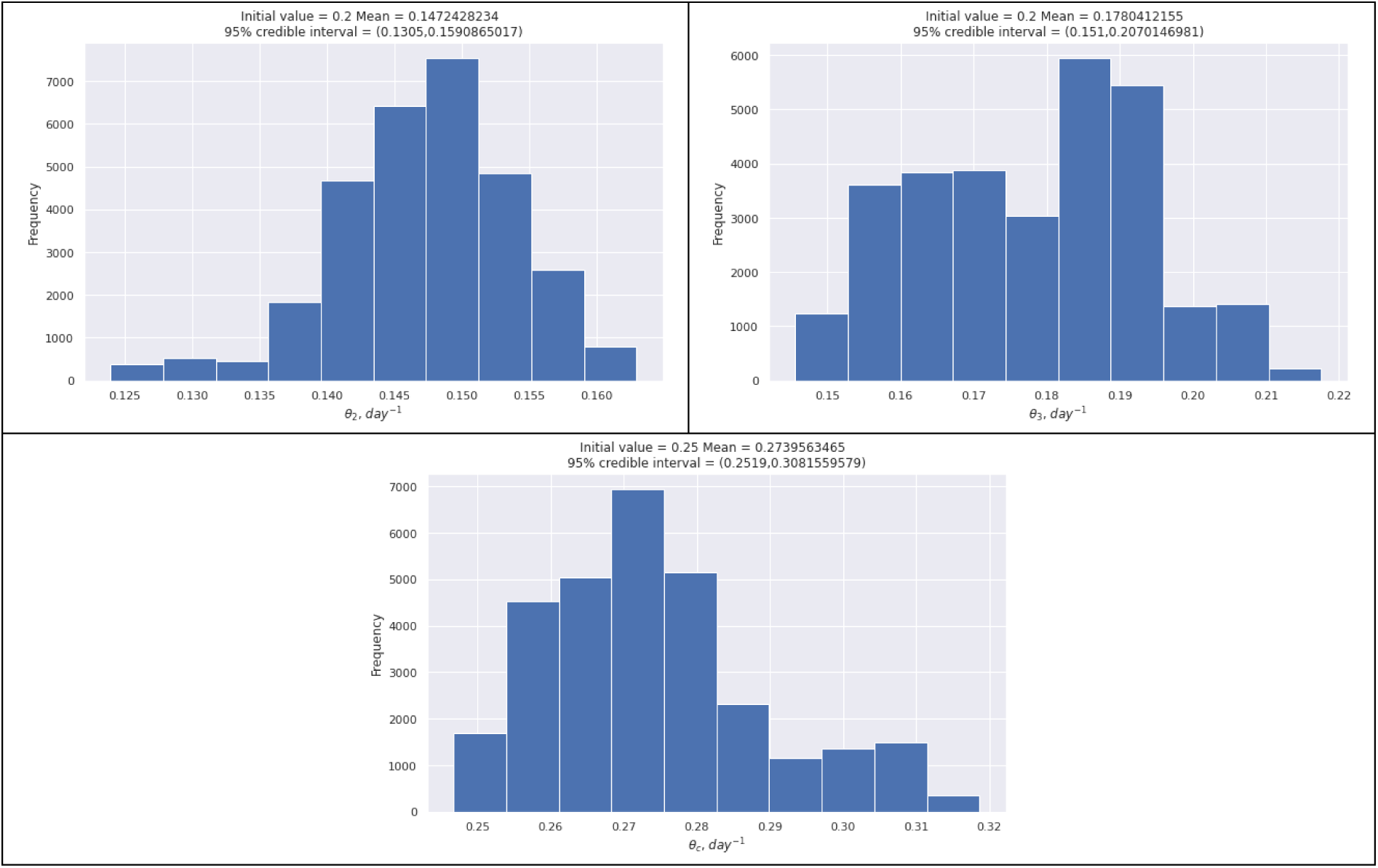
Histograms of the last 3×10^4^ states of the Markov chains for the model parameters

Despite the fact that some chains have not fully converged, due to low sensitivity and linear dependence of some parameters, the posterior distribution reaches convergence within few states, as demonstrated by figure 7. Therefore, an analysis of the sensitivity coefficients must be performed in the near future. The Integrated Autocorrelation Time (IACT) of the posterior was of the order of 100 states.

**Figure 7.**
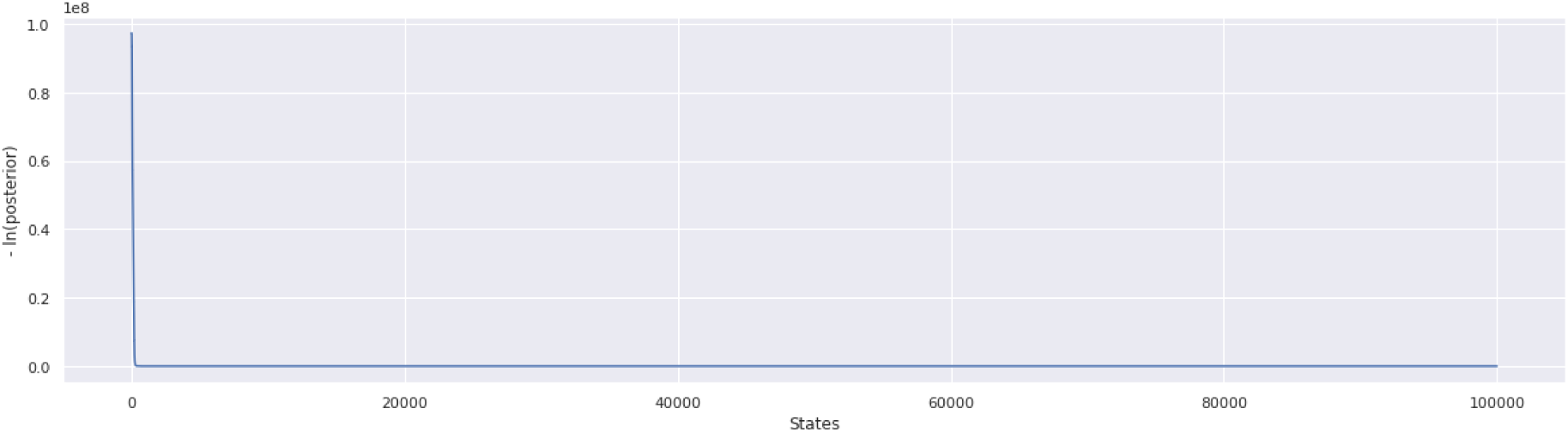
Variation of the posterior

## SIMULTANEOUS ESTIMATION OF STATE VARIABLES AND MODEL PARAMETERS WITH THE PARTICLE FILTER

The simultaneous estimation of state variables and model parameters was also performed in [12] for a model similar to the one used in this work. However, these authors augmented the vector of state variables with the parameters, which were modeled by random-walk processes. In the present work, the algorithm of Liu & West is used in order to avoid degeneration of the particles [5], which is more likely to occur with the algorithms used in [12]. The particle filter was applied with 5000 particles.

The state estimation problem was solved here with the data of the city of Rio de Janeiro [4], from February 28, 2020 to May 05, 2020. The means and standard deviations of the last 3×10^4^ states of the Markov chains were used for the approximate Gaussian distributions for the parameters at time t=0 in the particle filter.

The estimations obtained for the numbers of reported cases and reported deaths are shown in figures 8.a,b, respectively, together with the observed data. These figures reveal a significant improvement in the agreement between estimated and observed quantities, as compared to the solution of the parameter estimation problem with MCMC (see figures 4.b and 5.b). Such an improvement results from the fact that the parameters are now allowed to vary in time and they are dynamically estimated by the particle filter together with the state variables.

**Figure 8.**
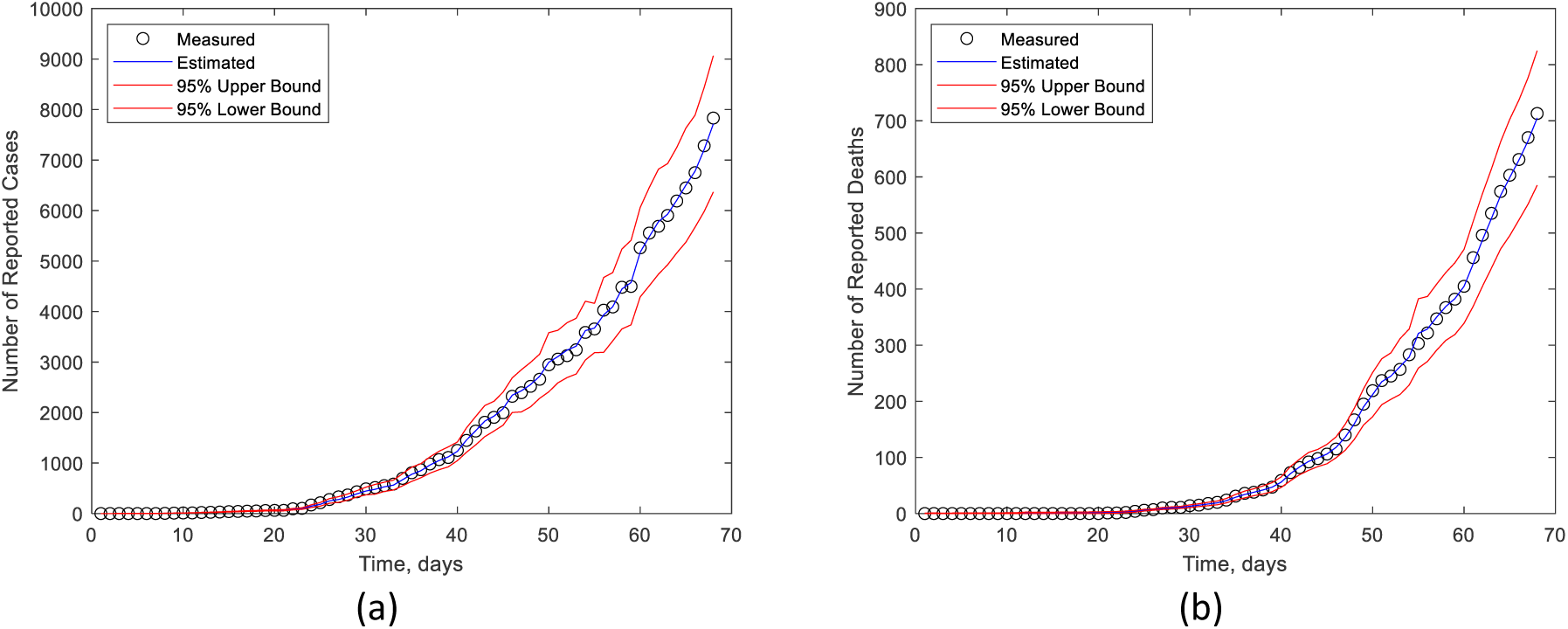
Comparison of estimated and observed numbers of reported cases (a) and reported deaths (b) obtained with the Particle Filter

The dynamic estimation of the model parameters is presented by figure 9. This figure shows a reduction of the transmission probability β_1_ at the same time that β_3_ increases, respectively for asymptomatic and hospitalized individuals. The transmission probability β_2_ for symptomatic individuals remains constant during the period. The rates of the disease progression σ_1_ and σ_2_ were not significantly modified and the initial means remain inside their 95% credibility intervals during the period, although they exhibit reduction and increase trends, respectively. While the rate σ_3_ decreases, σ_4_ increases, with behaviors analogous to σ_1_ and σ_2_, respectively. As for the probability of a symptomatic individual to be tested, a_2_, we observed a reduction trend, reaching a value around 0.065 at the end of the observed period. The other parameters were not significantly modified from their means, at the 95% credibility level.

**Figure 9.**
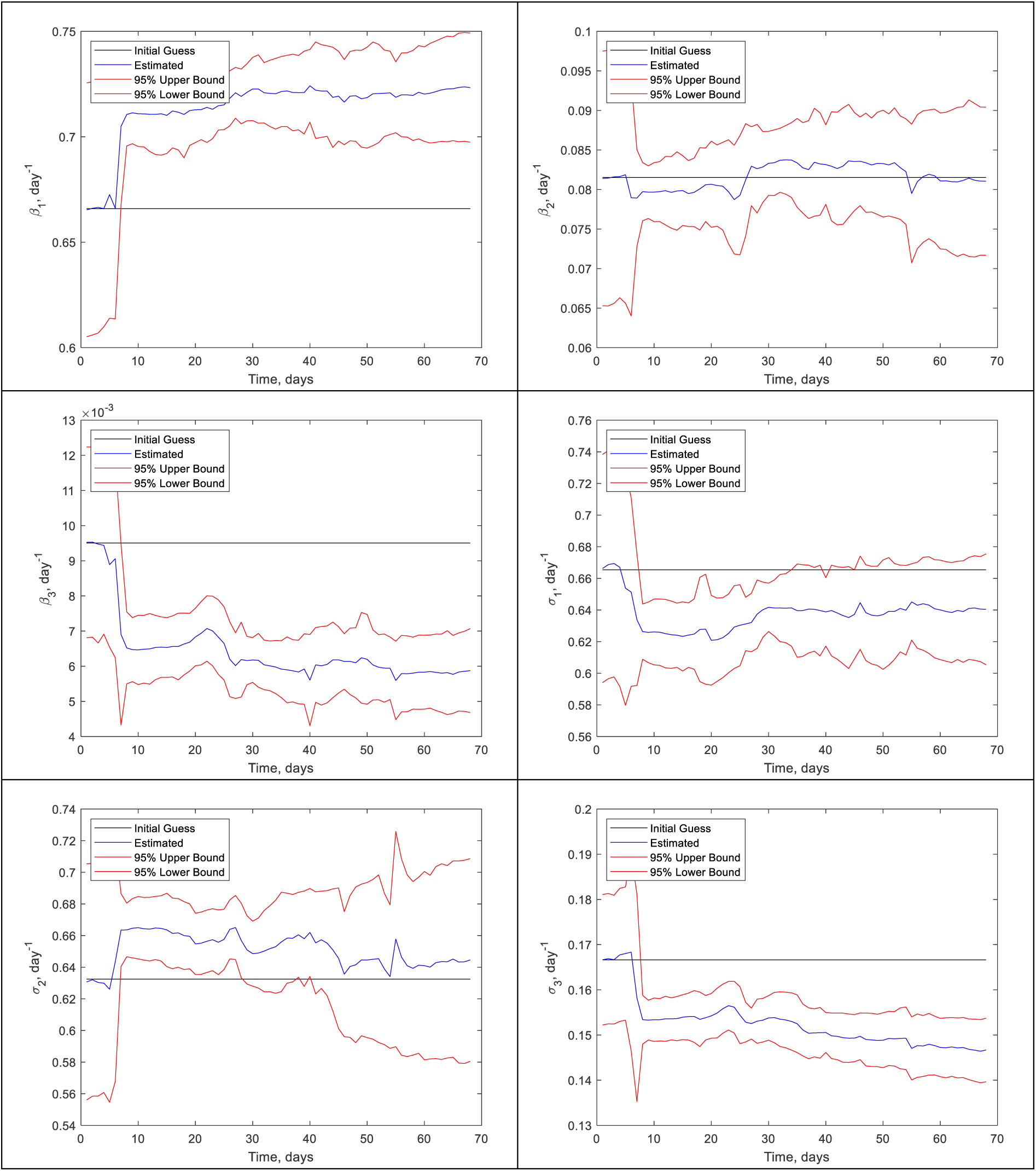
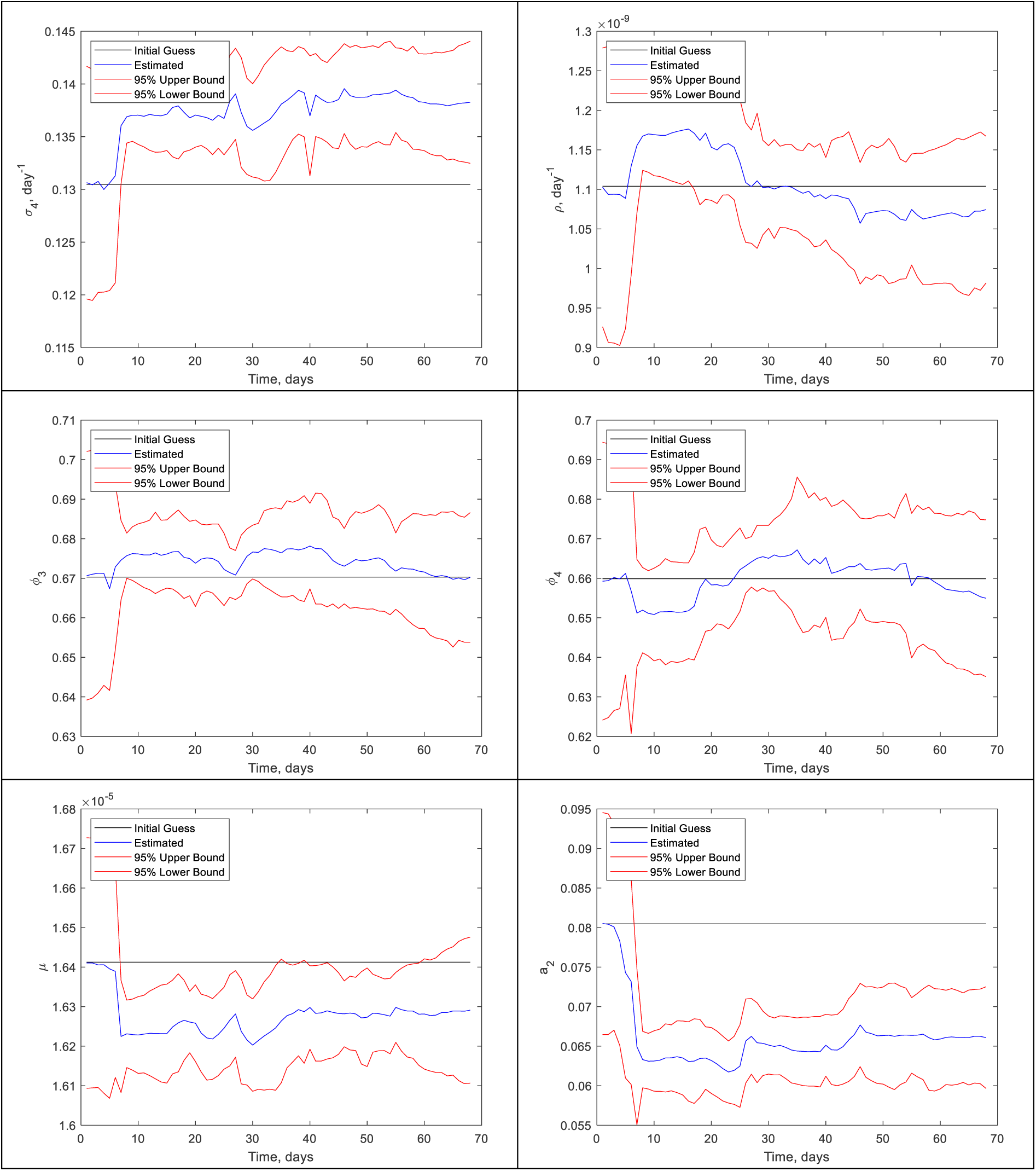
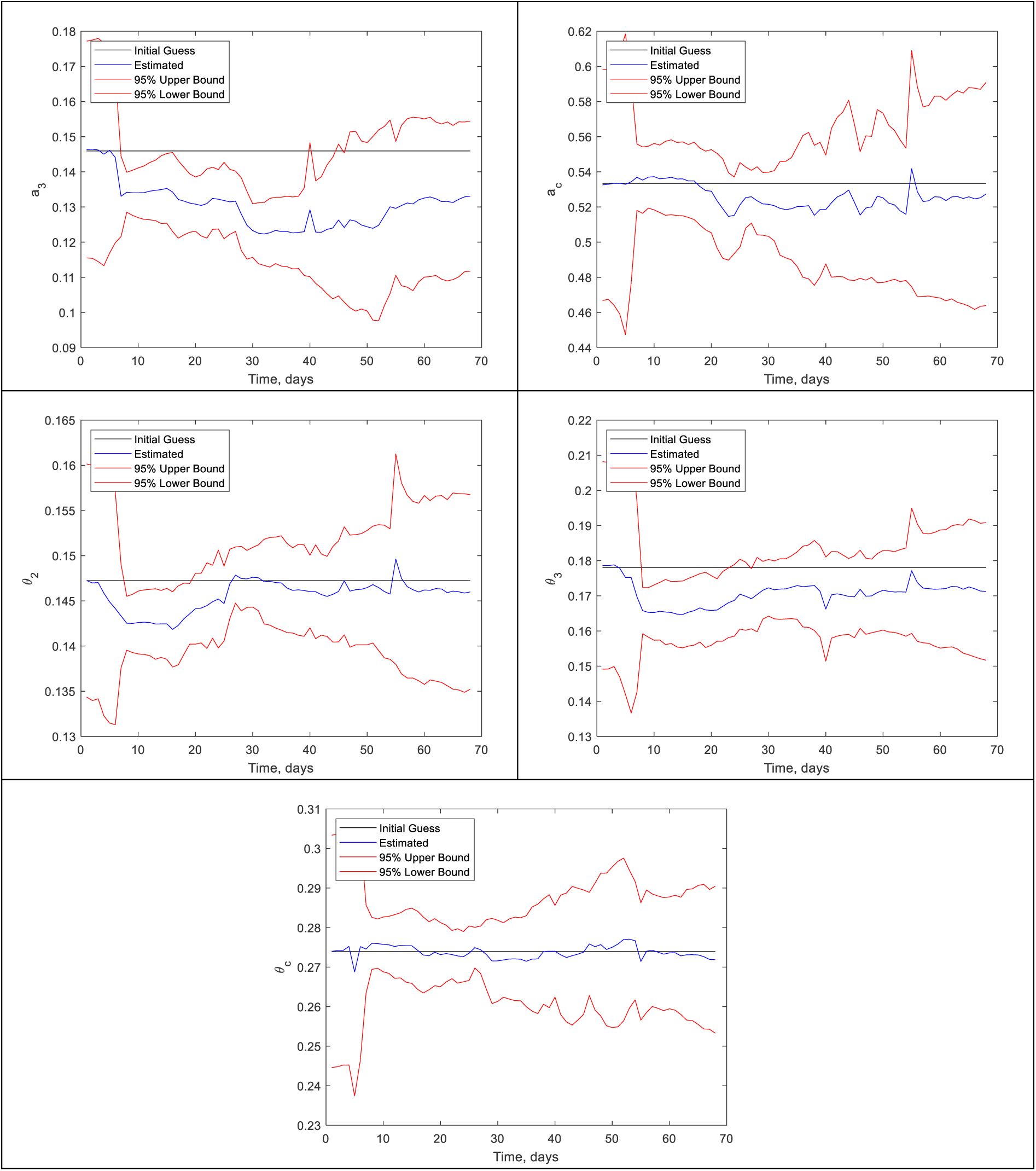
Dynamic estimation of the model parameters

Monte Carlo simulations were then made for 20 future days, starting on May 05, 2020, considering uncertainties in the model parameters and state variables. Initial conditions for the state variables and their corresponding distributions were those estimated with the particle filter for May 05, 2020. Distribution of the model parameters were also given by the estimations on this same date. Figures 10.a,b present the observed (shown by circles) number of cases and number of deaths, respectively, the estimated means and the 95% credibility intervals (given by blue and red lines, such as in figures 8.a,b), as well as the predictions obtained with the Monte Carlo simulations (yellow lines). These figures also show the median and the 2.5% and 97.5% quantiles of the Monte Carlo simulations (black lines), as well as the observed data from May 06, 2020 to May 15, 2020 (crosses). <u>Figures 10.a,b clearly show that the present model, with the parameters obtained with the Particle Filter, accurately fits the number of reported cases and the number of reported deaths, for 10 days ahead of the period used for the solution of the state estimation problem.</u> The observed number of reported cases tend follow the median line of the Monte Carlo simulations, with a trend towards the 2.5% quantile line for the last three days. On the other hand, the observed number of deaths follows very closely the 97.5% quantile line of the predictions obtained with the present model and correspondent estimated parameters.

**Figure 10.**
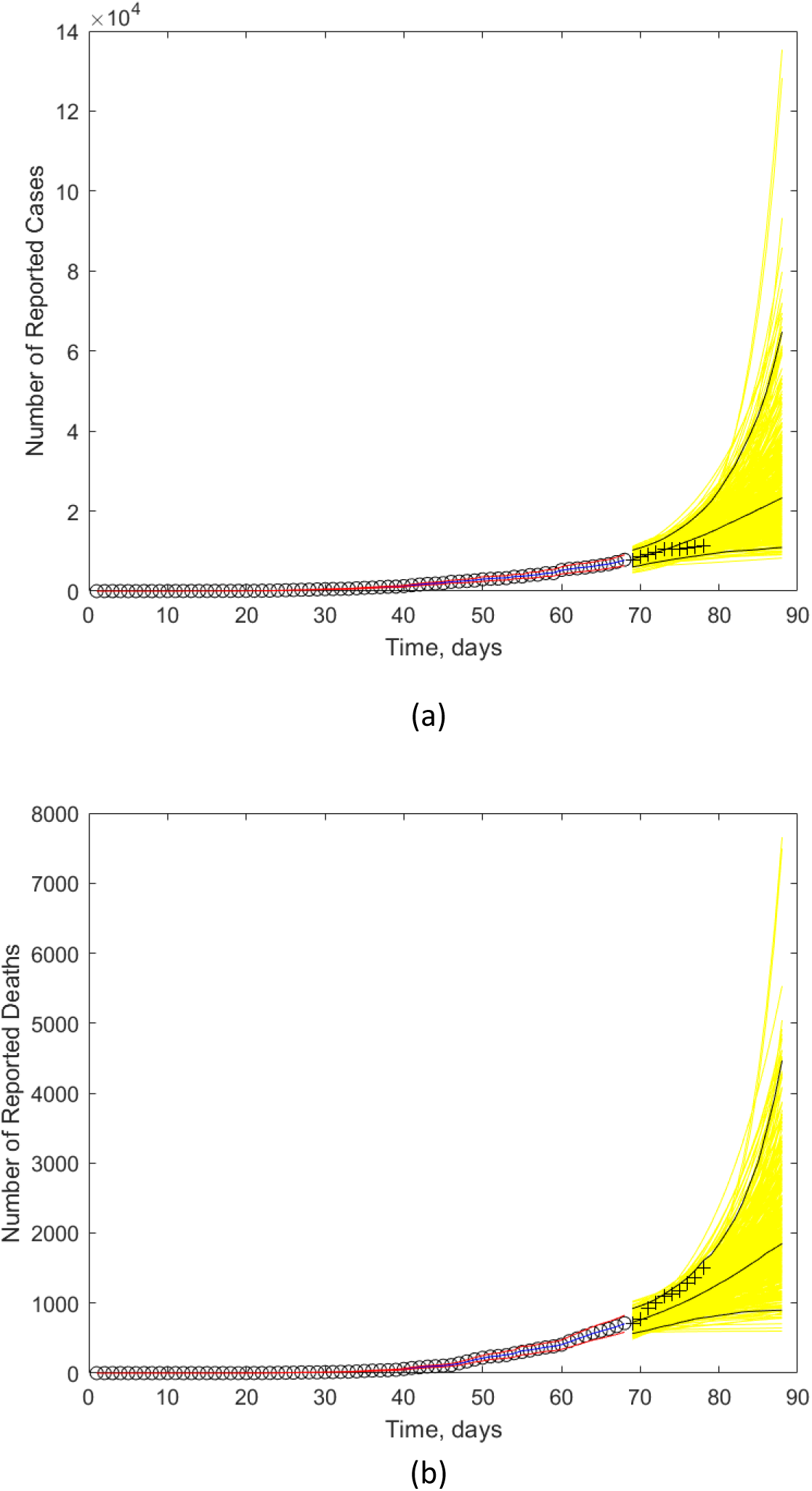
Comparison of the Monte Carlo simulations (yellow lines) and observed numbers of reported cases (a) and reported deaths (b). (Black lines represent the median and the 2.5% and 97.5% quantiles of the Monte Carlo simulations shown by the yellow lines. Circles indicate the observed data used for the solution of the state estimation problem. Crosses indicate the observed data used for the validation of the model from May 06, 2020 to May 15, 2020. Blue and red lines represent the mean and the 95% credibility interval of the solution of the state estimation problem. **The yellow curves starting on May 06, 2020 and finishing on May 25 are values predicted by the model**.)

Figures 10.a,b suggest that the <u>particle filter must be used to periodically update the estimation of state variables and model parameters, so that future predictions (in a period of about 10 days) can be made.</u> The Monte Carlo predictions for other state variables are presented by figure 11.

**Figure 11.**
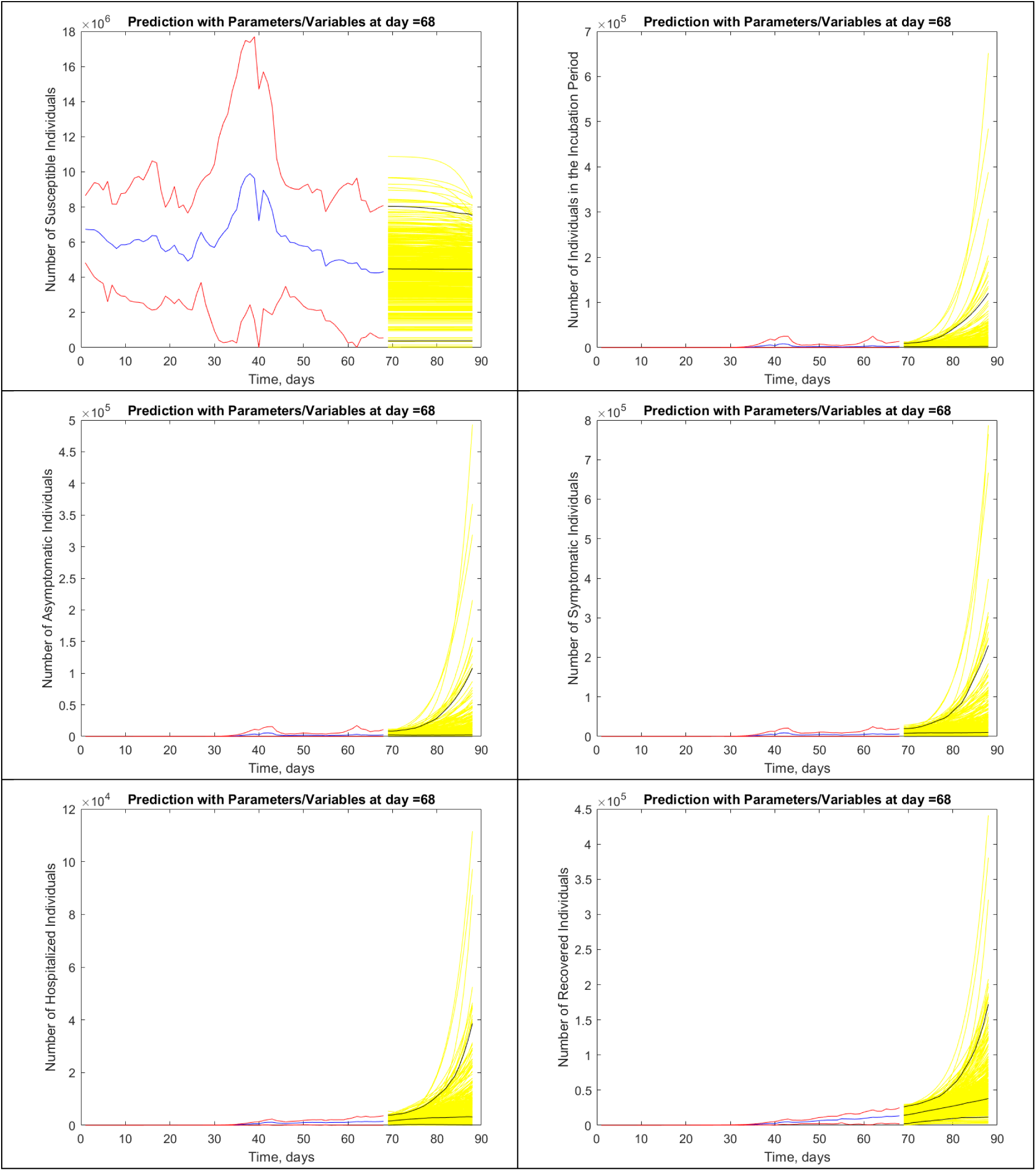
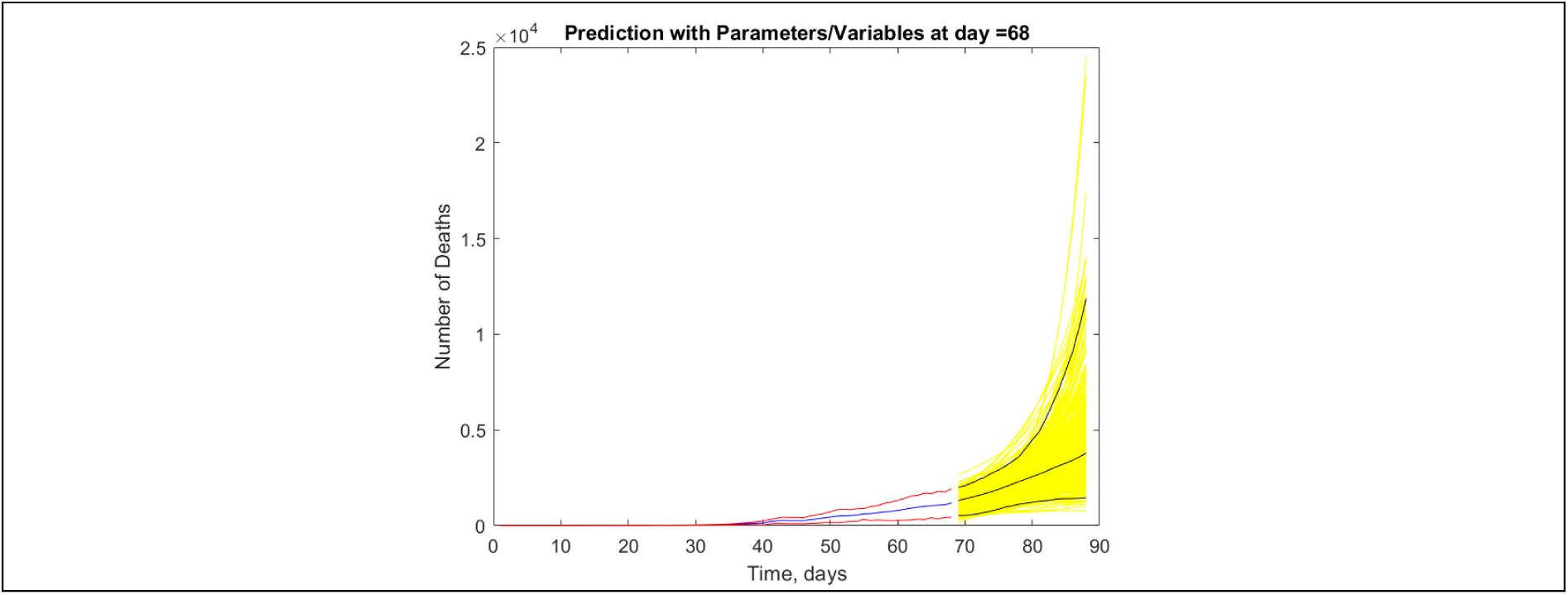
Monte Carlo predictions (yellow lines) for other state variables (Black lines represent the median and the 2.5% and 97.5% quantiles of the Monte Carlo simulations shown by the yellow lines. Circles indicate the observed data used for the solution of the state estimation problem. Crosses indicate the observed data used for the validation. Blue and red lines represent the mean and the 95% credibility interval of the solution of the state estimation problem.). **The yellow curves starting on May 06, 2020 and finishing on May 25 are values predicted by the model.)**

Figures 12-15 respectively present the ratio of infected individuals per reported cases, the indexes of under-reported cases and deaths, and the effective reproduction number, which were estimated during the period used for the solution of the state estimation problem with the particle filter, that is, from February 28, 2020 to May 05, 2020.

**Figure 12.**
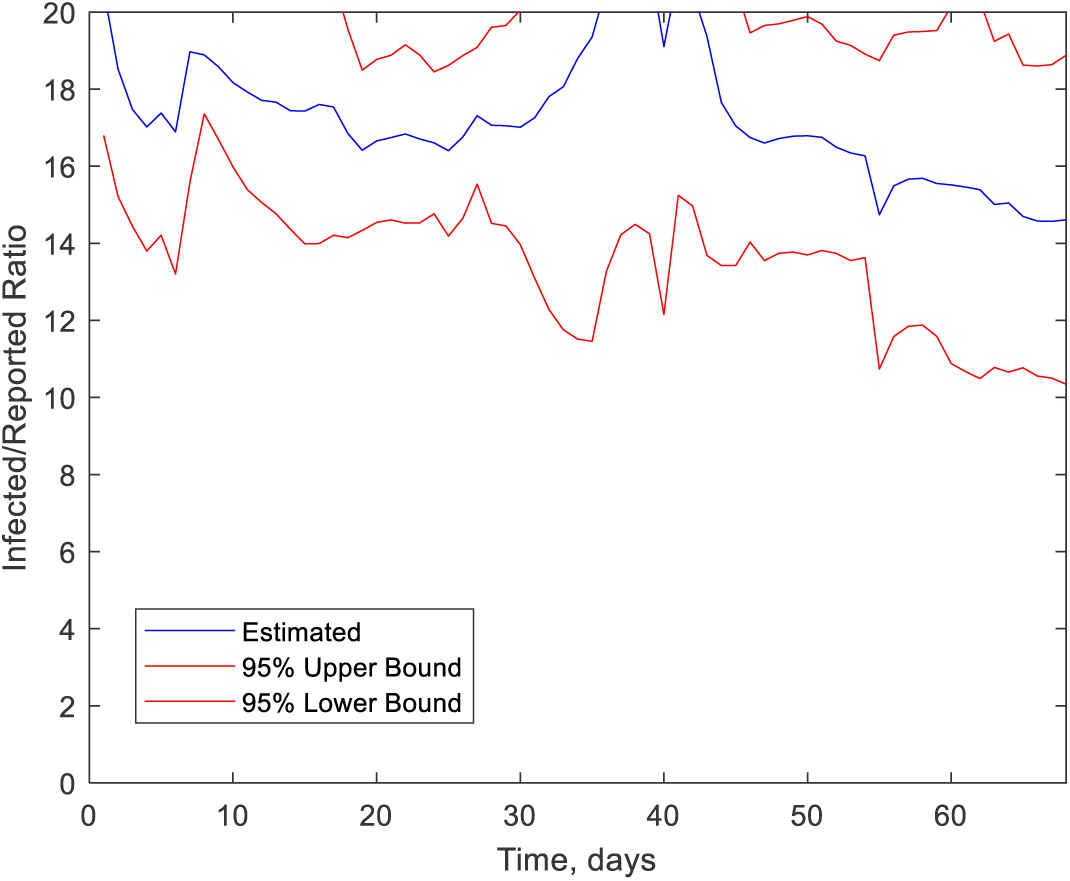
Ratio of Infected Individuals per Reported Cases

**Figure 13.**
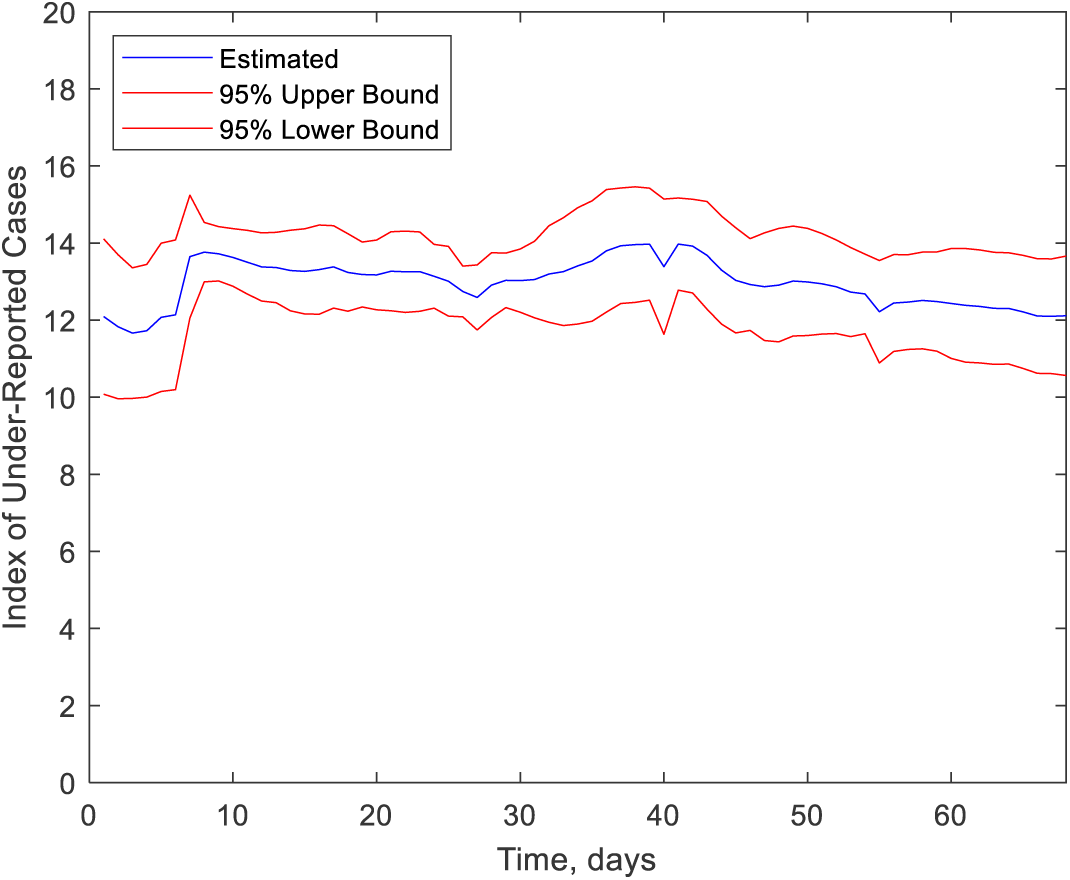
Index of Under-Reported Cases

**Figure 14.**
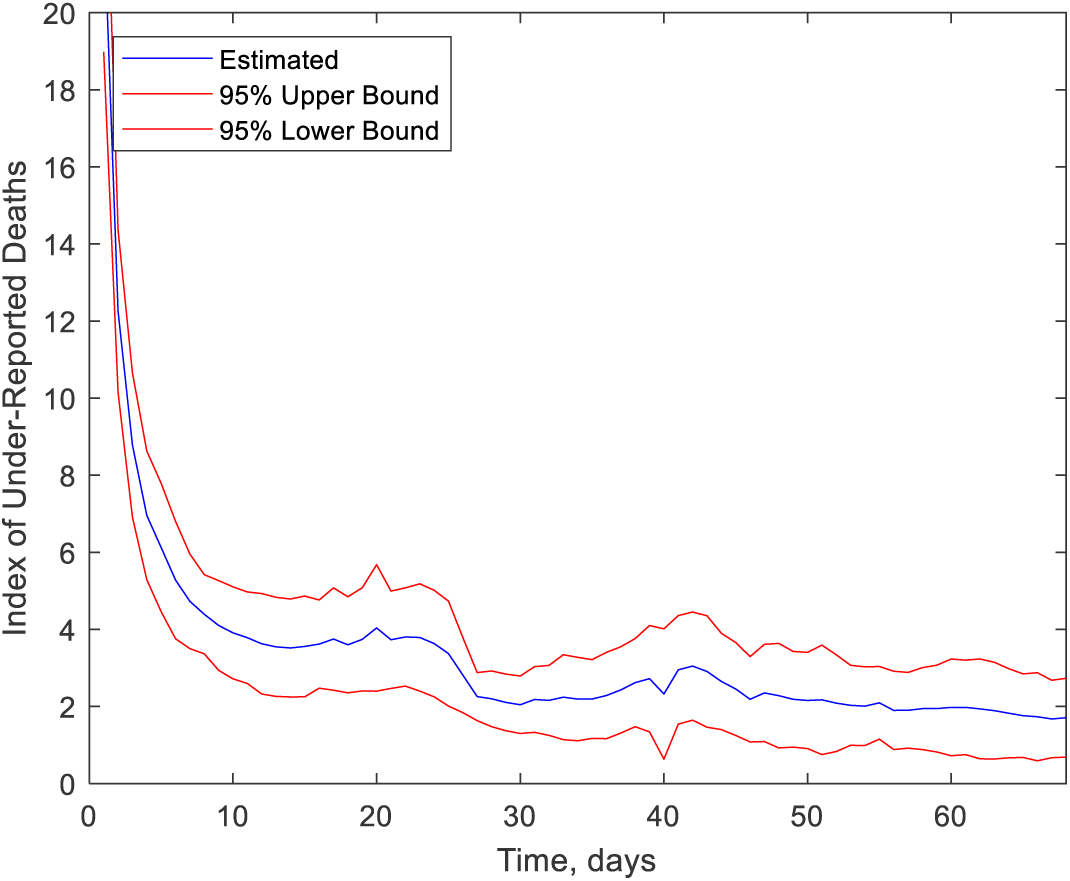
Index of Under-Reported Deaths

**Figure 15.**
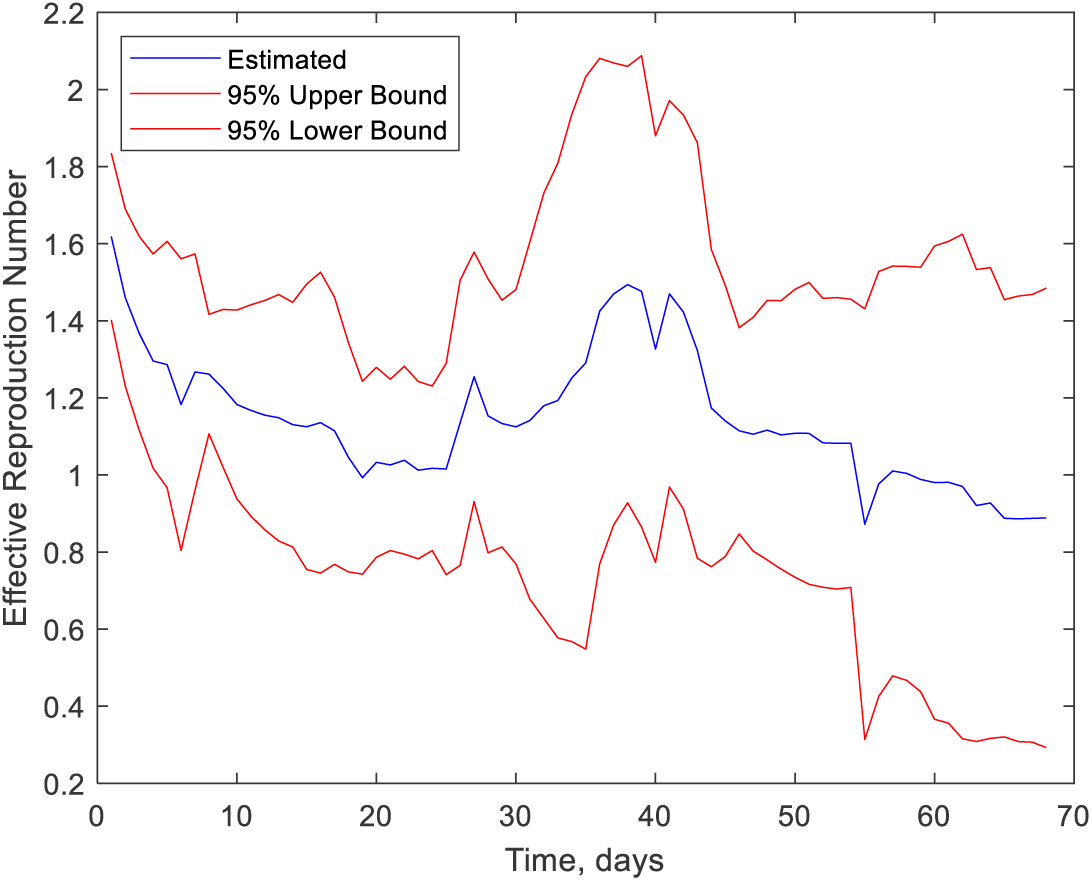
Effective Reproduction Number

The *Ratio of Infected Individuals per Reported Cases* is given by:

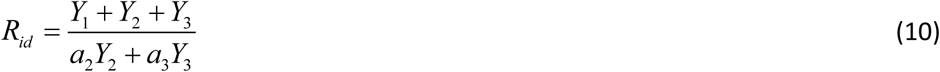

Therefore, this ratio includes not only tested individuals, but also the asymptomatic individuals that are not tested. The mean of this ratio is around 15 on May 05, 2020.

Different from the above ratio, the *Indexes of Under-Reported Cases and Deaths* only involve tested individuals and are given, respectively, by:

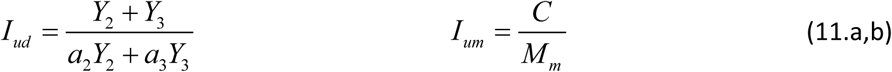

The means of the above indexes on May 05, 2020 were estimated around 12 and 2, respectively. The *Effective Reproduction Number* for the disease was calculated with:

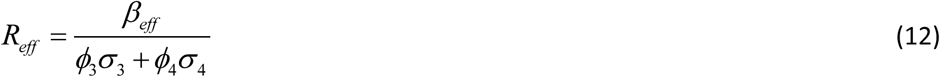

where the effective transmission probability that gives the force of the infection is given by:

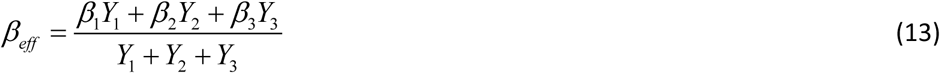

The mean value of the effective reproduction number was around 1.6 on February 28, 2020 and dropped to around 0.9 on May 05, 2020. However, uncertainties related to this parameter are large and the effective reproduction number is between 0.3 and 1.5, at the 95% credibility level.

## Data Availability

Only public data has been used, as referenced in the manuscript.

## Notes

### Competing Interest Statement

The authors have declared no competing interest.

### Funding Statement

CNPq
CAPES
FAPERJ

### Author Declarations

Only public data available online has been used, as referenced in the manuscript.

## REFERENCES

[1] Hernández, A., Dulikravich, G., Blower, S., Colaço, M., Moral, R., Identification of Parameters in a System of Differential Equations Modeling Evolution of Infectious Diseases, Proceedings of 2008 ASME International Design Engineering Technical Conferences & Computers and Information in Engineering Conference, IDETC/CIE 2008, August 3–6, 2008, Brooklyn, New York, USA.

[2] Gamerman, D. and Lopes, H.F., 2006, Markov Chain Monte Carlo: Stochastic Simulation for Bayesian Inference, Chapman & Hall/CRC, 2nd edition, Boca Raton.

[3] Kaipio, J. and Somersalo, E., 2004, Statistical and Computational Inverse Problems, Applied Mathematical Sciences 160, Springer-Verlag.

[4] http://cor.rio/, consulted on April 30, 2020.

[5] Liu, J., West, M., 2001. Combined parameter and state estimation in simulation based filtering, in: A. Doucet, N. de Freitas, and N. Gordon, (Eds.), New York, S. (Ed.), Sequential Monte Carlo Methods in Practice. pp. 197–217.

[6] Neil Ferguson et al, Impact of non-pharmaceutical interventions (NPIs) to reduce COVID-19 mortality and healthcare demand, 16 March 2020, Imperial College COVID-19 Response Team.

[7] S. Cauchemez, F. Carrat, C. Viboud, A. J. Valleron and P. Y. Boelle, A Bayesian MCMC approach to study transmission of inuenza: application to household longitudinal data, Statist. Med. 2004; 23: 3469–3487.

[8] Cotta, Renato Machado, and Carolina Palma Naveira-Cotta. Modelling the COVID-19 epidemics in Brasil: Parametric identification and public health measures influence. medRxiv (2020).

[10] Timothy W Russell, Joel Hellewell, Sam Abbott, Nick Golding, Hamish Gibbs, Christopher I Jarvis, Kevin van Zandvoort, CMMID nCov working group, Stefan Flasche, Rosalind M Eggo, W John Edmunds & Adam J Kucharski, Using a delay-adjusted case fatality ratio to estimate under-reporting, https://cmmid.github.io/topics/covid19/global_cfr_estimates.html, consulted on May 13, 2020.

[11] https://brasilemsintese.ibge.gov.br/populacao/taxas-brutas-de-mortalidade.html, consulted on April 10, 2020.

[12] Dureau, Joseph, Konstantinos Kalogeropoulos, and Marc Baguelin. “Capturing the time-varying drivers of an epidemic using stochastic dynamical systems.” Biostatistics 14.3 (2013): 541–555.

[13] Linka, Kevin, et al. “Outbreak dynamics of COVID-19 in Europe and the effect of travel restrictions.” Computer Methods in Biomechanics and Biomedical Engineering (2020): 1–8.

[14] Peirlinck, Mathias, et al. “Outbreak dynamics of COVID-19 in China and the United States.” Biomechanics and Modeling in Mechanobiology (2020): 1.

